# Assembling the Challenge of Multimorbidity in Zimbabwe: A Participatory Ethnographic Study

**DOI:** 10.1101/2024.08.06.24311557

**Authors:** Justin Dixon, Efison Dhodho, Fionah Mundoga, Karen Webb, Pugie Chimberengwa, Trudy Mhlanga, Tatenda Nhapi, Theonevus Tinashe Chinyanga, Justice Mudavanhu, Lee Nkala, Ronald Nyabereka, Gwati Gwati, Gerald Shambira, Trust Zaranyika, Clare I.R. Chandler, Rashida A. Ferrand, Chiratidzo Ellen Ndhlovu

## Abstract

Multimorbidity, increasingly recognised as a global health challenge, has recently emerged on the health agendas of many lower-income countries, including in Africa. Yet with its conceptual origins in the global North, its meaning and possible utility for stakeholders in lower-resources settings remains abstract. This study drew together policymakers, public health practitioners, academics, health informaticians, health professionals, and people living with multimorbidity (PLWMM) in Zimbabwe to understand: What is the transformative potential and possible limitations of elevating multimorbidity as a priority in this setting? To bring these different perspectives into conversation, we used a participatory ethnographic design that involved a health facility survey, participant-observation, in-depth interviews, audio-visual diaries, and participatory workshops. Multimorbidity, we found, was new to many respondents but generally viewed as a meaningful and useful concept. It foregrounded a range of challenges related to the ‘vertical’ organisation and uneven funding of different diseases, while revealing promising opportunities for integration across entrenched silos of knowledge and practice. However, with capacity and momentum to address multimorbidity currently concentrated within the HIV programme, there was concern that multimorbidity could itself become verticalized, undercutting its transformative potential. Participants agreed that responding to multimorbidity requires a decisive shift from vertical, disease-centred programming to restore the comprehensive primary care that undergirded Zimbabwe’s once-renowned health system. It also means building a policy-enabling environment that values generalist (as well as specialist) knowledge, ground-level experience, and inclusive stakeholder engagement. The ‘learning’ health system, we conclude, represents a promising conceptual lens for unifying these imperatives, providing a tangible framework for how knowledge, policy, and practice synergise within more self-reliant, person-centred health systems able to respond to ever-evolving complex health challenges like multimorbidity.

## 1. Introduction

Multimorbidity, commonly defined as the co-occurrence of two-or-more long-term conditions in one individual, has been described as the next ‘global pandemic’.[1] In countries undergoing epidemiological transitions, including in Africa, multimorbidity has been characterised as a ‘clash’ of persisting communicable diseases (notably HIV and tuberculosis (TB)) and rising non-communicable diseases (NCDs) that require multiple forms of expertise and coordinated care to manage.[2] Yet, health systems remain largely organised around specialist rather than generalist knowledge,[3] which in many African nations translates into siloed organisation of care, fuelled by ‘vertical’ single-disease programming.[2] Priority-setting initiatives for multimorbidity in a global context[4–6] including in sub-Saharan Africa specifically[5,6] highlight the need to identify common disease ‘clusters’ and shared determinants; improve multimorbidity prevention and treatment; and more broadly restructure health systems to become more integrated and person-centred. The COVID-19 pandemic has since highlighted the importance of prioritising multimorbidity, with the virus disproportionately affecting those with multiple ‘underlying conditions’.[7]

The social sciences, including medical anthropology, have been integral in advancing understandings of multimorbidity. Qualitative research in a range of low-resource contexts including in Africa has shed light on the systemic vulnerabilities that expose marginalised people and populations to multimorbidity,[8,9] the way that health and social challenges cumulatively intertwine to coproduce and exacerbate multimorbidity,[10,11] and the challenges faced by patients and providers navigating fragmented health systems designed for single disease care.[12–15] Qualitative studies have also contributed to recognition that frameworks to integrate care are extremely challenging to implement in practice and often do not go far enough to tackle multimorbidity’s social and structural determinants. Bosire et al,[12] for instance, demonstrated that South Africa’s Integrated Chronic Disease Management (ICDM) Model, designed to provide integrated person-centred care for long-term conditions, has faced considerable challenges delivering on its intended outcomes due to resource scarcity, partial implementation, continued fragmentation, and emphasis on pharmaceutical disease care at the expense of patients’ social and economic context.

With multimorbidity gaining traction as a global health priority, a growing number of studies have critically interrogated what it is currently ‘doing’ as a concept and whether it is proving as radical and transformative as it has been hoped to be.[16–18] On the one hand, multimorbidity has become a focal point for tensions that have been long building across the spectrum of medicine and global health, sparking cross-cutting recognition of the evident breakdown of ‘vertical’ funding and governance mechanisms, parallel research and data infrastructures, the proliferation of clinical sub-specialities and over generalism and public health; and fragmented, unevenly resourced care delivery systems.[17] As such, multimorbidity may be a powerful concept for confronting these systemic issues and realigning of health systems with increasingly complex health needs. At the same time, multimorbidity is evidently a disease-centred construct and emergent of the very Euro-Western philosophical traditions that have made the single disease model so intractable.[16–18] Recognition of this has prompted several initiatives – including within the current Collection – to reconceptualise multimorbidity to put people-in-context, rather than diseases, back at its theoretical centre.[16,19,20] Yet with little empirical research into the meaning and significance of multimorbidity within particular health system contexts, particularly in low-resource settings, these conversations remain abstract. Inclusive, bottom-up understandings of multimorbidity are required to ensure that multimorbidity is able to fulfil some of the promises that have been pinned to it and that it does not inadvertently perpetuate disease-centred care.[17]

This article presents findings from a collaborative anthropological study that sought to address the question: what is the transformative potential and possible limitations of elevating multimorbidity as a priority in Zimbabwe? To answer this question we brought together a diversity of perspectives on multimorbidity from across the country’s health and academic sectors to better understand the tensions, challenges, and opportunities that multimorbidity brought to the fore for differently-situated respondents and to identify potential benefits and limitations of multimorbidity as a conceptual lens and organising principle. Because multimorbidity in Zimbabwe, like many African nations, was only just starting to emerge on the radar and was not yet part of a formal strategy, this was an opportune moment to develop a formative agenda and set of priorities with already-well positioned respondents while creating new partnerships and relationships to take these forward. The aim was to engage stakeholders in a way that was holistic, critical, and commensurate across different disciplines, fields, and perspectives, and would therefore help ensure that transformative potentials of multimorbidity were optimised while minimising any possible limitations or harms.

## 2. Study Setting and Design

This article is based on findings from the KnowM^M^ study (2021-2024), an interdisciplinary global health research collaboration centred on a participatory ethnographic study of multimorbidity within the Zimbabwean health system.

### 2.1. Study setting

Zimbabwe is a lower-middle-income country with a population of 16.3 million.[21] Following independence in 1980, huge strides were made in expanding access to healthcare, including in moving from an urban, curative, and racially-biased health system to one focused on delivering affordable primary healthcare to underserved communities.[22–24] Zimbabwe came to boast one of Africa’s strongest health systems, with thriving teaching hospitals and academic sector, well-trained health workforce, and decentralised care system organised rationally around its essential medicine list and national treatment guideline (EDLIZ) and co-produced training manuals.[25] However, the achievements of the 1980s–90s were undone by political instability, economic structural adjustment (which decreased public spending in favour of privatisation), hyperinflation, the reintroduction of user fees in hospitals and urban clinics, and the HIV and AIDS epidemic.[26] Since then, Zimbabwe experienced among the steepest exponential rises in NCDs in sub-Saharan Africa,[27] and modelling suggests that multimorbidity, particularly among people living with HIV, will rise sharply by 2035.[28] The health system remains fragmented, under-resourced and oriented towards infectious disease care. It has also experienced high rates of health workers attrition and regular collapses over the last two decades, most recently during the COVID-19 pandemic.[29]

KnowM^M^ was conducted in four provinces of Zimbabwe: Harare, Bulawayo, Mashonaland East, and Matabeleland South (Figure 1). Harare and Bulawayo are metropolitan provinces, with the majority of political institutions, universities, and all Central hospitals. They also both share a similar provincial health system structure, with primary healthcare for the majority provided by City Councils and most referrals going straight to one of the Central hospitals. Both also have large private sectors made up of primary and secondary facilities (for the affluent minority), as well as private pharmacies and laboratories. Mashonaland East, bordering Harare, is a predominantly rural province in which most public healthcare is run by the central government, which includes primary, secondary, and tertiary care, with referrals for quaternary care generally sent to Harare. Matabeleland South is a predominantly rural province with a broadly similar structure to Mashonaland East, with quaternary referrals referred mainly to Bulawayo rather than to Harare. The four provinces were selected to capture urban and rural settings, a balance of provinces in the majority Mashonaland region and minority Matabeleland region, as well as to capture the main differences in health system structure and referral relationships.

**Figure 1:**
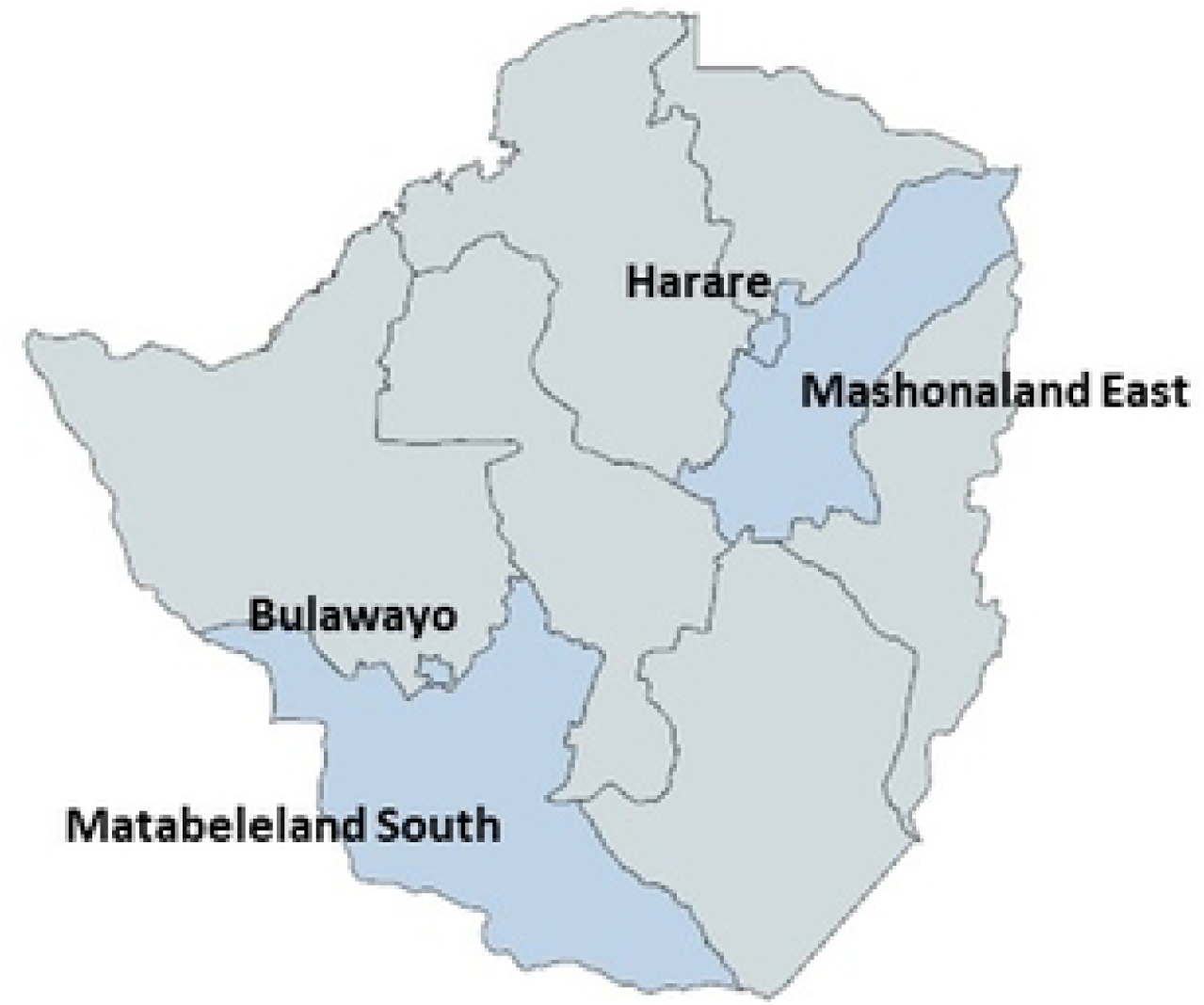
Provinces of Zimbabwe included in the KnowM^M^ study.

### 2.2. Study design

KnowM^M^ used a participatory ethnographic study design. Informed by mapping work of the emerging field of global multimorbidity research,[6,17] primary data collection including participant recruitment took place in Zimbabwe between 01-09-2022 and 31-12-2023 by an interdisciplinary team of medical anthropologists (JD, FM, CC), clinical academics (RAF, GS, TZ, CEN), and public health practitioners from project implementing partner OPHID (KW, ED, PC, TM, TN, and TTC), supported by key Directorates within the Ministry of Health and Child Care (MoHCC) (RN, JM, LN, GG). Our study design is informed by the principle of “slow co-production”[30] which includes participants not just as sources of data but as active agents in the production of knowledge. This was important given that the concept of multimorbidity has emerged from high-income settings, with uncertainty about its relevance beyond the rich world. In engaging with participants, we invited them to think ‘with’ the concept of multimorbidity – where necessary, providing the working definition of ‘multiple long-term conditions’ – but critically, without reifying it as a category and allowing participants to drive conversations about its meaning and significance in the Zimbabwean context.

In total, 130 individuals took part in the study (overview of participants provided in Table 1). Data collection commenced with a survey of health facilities (n=30) in the four provinces, purposively selected to provide a broad, formative understanding of the capacity of facilities at different levels of care in different provinces for addressing NCDs and multimorbidity (see Table 2). Questions pertained to services and staffing; routines and challenges regarding the management of NCDs and multimorbidity; and preparedness for managing specific NCDs (hypertension, diabetes, chronic respiratory disease, and depression), the latter based on WHO facility indicators.[31] Surveys were administered with facility managers, with input on specific questions by clinicians, pharmacists, human resources, and health information officers. To gain a deeper appreciation of how multimorbidity was being engaged with in practice, we conducted participant-observation and in-depth interviews in 10 facilities from the survey purposively selected to represent different levels of care. Participants were generally nurses or physicians (depending on level of care) to whom we were referred by facility managers, with expertise in general medicine, infectious diseases (especially HIV and TB), physical NCDs (including cardiometabolic conditions, oncology, and rheumatology), and mental health conditions (see Table 1). Participant-observation typically took place over 1-3 days and involved ‘shadowing’ participants to understand practices and routines related to multimorbidity, during which fieldnotes were taken on encrypted tablets. Interviews were conducted using bespoke interview guides developed from a set of high-level pre-determined themes as well as findings from observation, and were conducted where possible at participants’ places of work during quiet periods. Interviews lasted approximately 1 hour and were audio-recorded unless participants declined, in which case detailed notes were taken instead. No repeat interviews were conducted. To gain a patient perspective, we included 23 PLWMM from facilities involved in participant-observation, purposively selected in consultation with attending health workers to represent differences in age, sex, geographical location, and conditions (<u>S1 Appendix</u>). We conducted in-depth interviews with all 23 PLWMM to capture illness and treatment-seeking narratives, understandings of their medical conditions, and experiences accessing care. We asked every third participant to keep 7-day audio-visual diary on encrypted smartphones to gain a deeper understanding of their experiences and routines. Where appropriate (e.g., for elderly patients), family members and carers assisted in the recording of the audio-visual diaries.

**Table 1:**
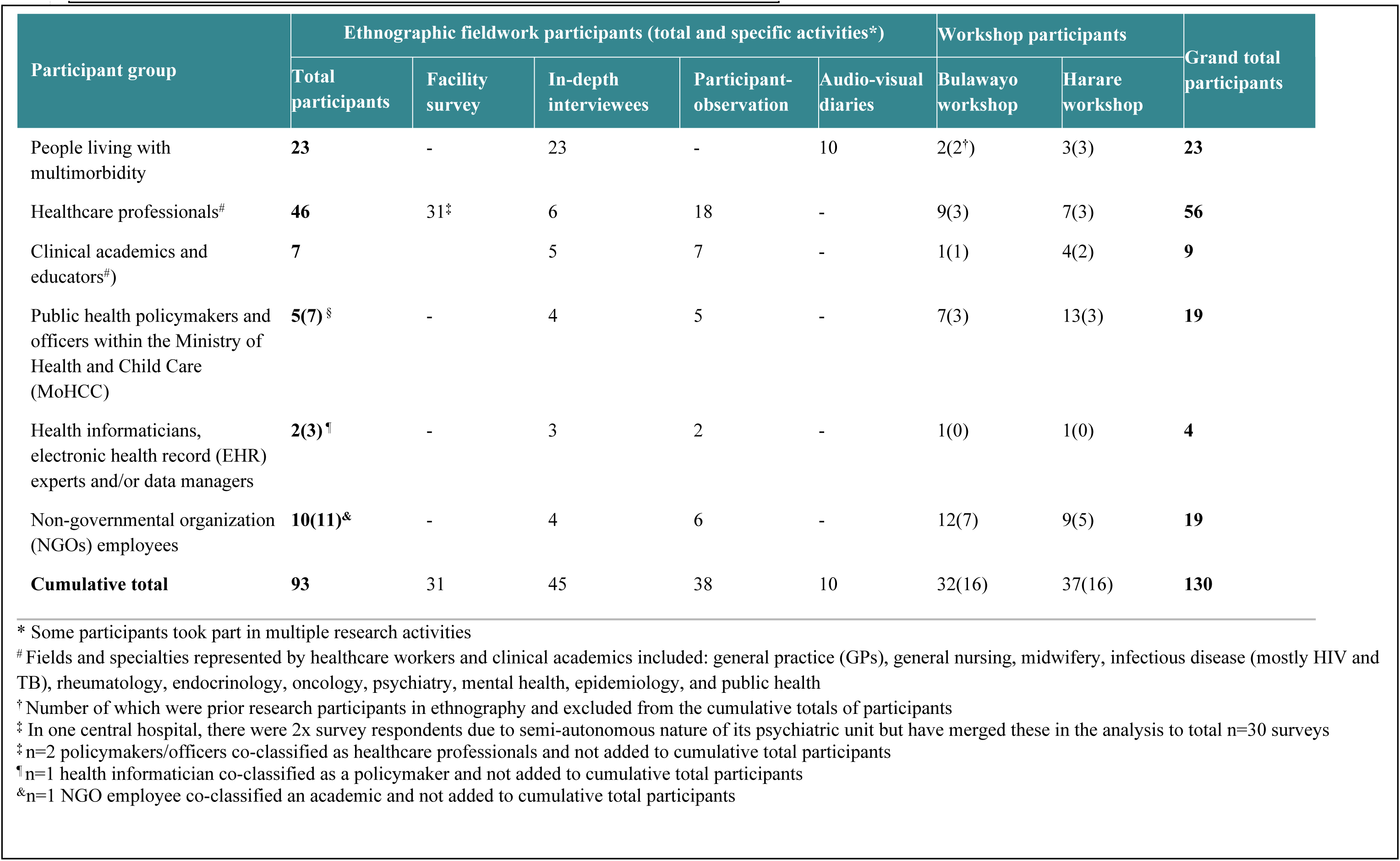
Participant demographics and activities breakdown.

**Table 2:**
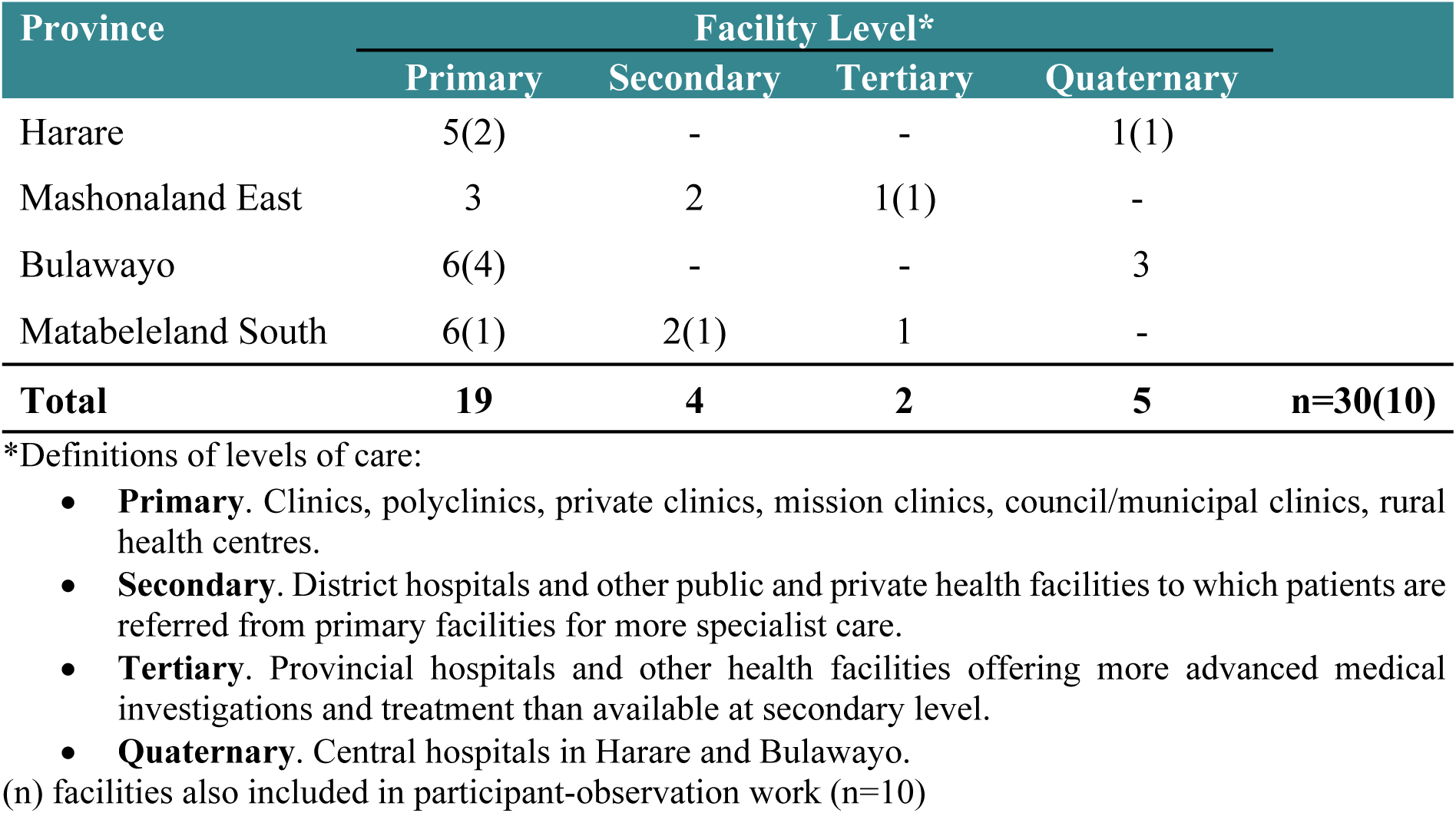
Health facility survey details.

To capture how multimorbidity was being engaged in other spaces within the health system beyond the service delivery level, we conducted in-depth interviews and, where appropriate, participant-observation with professionals working across Zimbabwe’s health and academic sectors. This included public health policymakers and practitioners in the MoHCC (n=7), health informaticians, electronic health record (EHR) experts and/or data managers (n=3), technical partners and non-governmental organisations (NGOs) (n=11), and academics/researchers at tertiary education and research institutions (n=7). Most of the latter were also at the top of their fields in their respective specialities, providing a clinical academic perspective; most were also lecturers, thus also offering a medical education perspective. Interviews and participant-observation were conducted using the same approach, timeframe, and data collection tools to those used at service delivery level, with some overlap given the aforementioned porosity of academic and clinical roles. Finally, we held two participatory workshops, one in Bulawayo (including participants and stakeholders from Bulawayo and Matabeleland South, n=32), and one in Harare (including participants and stakeholders from Harare and Mashonaland East, n=37). The aim was to collaboratively interpret primary data and co-develop a formative agenda comprised of priority areas, specific recommendations, and focal institutions/departments for taking forward. Participants were invited to bring their knowledge and experiences to bear on discussions, but in the process to reflexively situate and potentially challenge their own assumptions through engagement with others’ perspectives.

### 2.3 Data Analysis

All data sources were entered into NVivo 14 for analysis. Data was analysed using the principles of grounded theory – an inductive, theory-generating methodology that produces contextualised, process-oriented theories of social phenomena.[32] Analysis was performed on an ongoing, iterative basis by JD and FM that overlapped with, and was fed back into, the ongoing collection of data. During primary data collection, observational, interview, audio-video were coded, firstly through initial mapping and categorisation of data, before identifying key themes emerging from different perspectives. During the collaborative workshops, participants actively engaged in interpreting the findings and played a crucial role in the validation and refinement of emerging themes. Workshop participants further worked together to translate findings into a provisional multimorbidity agenda, which was subsequently refined and finalised during a final round of data analysis and feedback.

### 2.4. Ethical Considerations

Ethical approval was sought from the Medical Research Council of Zimbabwe (MRCZ/A/2842), Joint Research Ethics Committee of the Parirenyatwa Group of Hospitals and University of Zimbabwe Faculty of Medicine and Health Sciences (386/2021), the City of Harare, and the London School of Hygiene and Tropical Medicine (26469). All participants provided written informed consent to take part. Where appropriate, group briefings were conducted at healthcare facilities prior to asking individuals for consent. Before participant-observation, timeframes were agreed upon with participants such that they were always comfortable with the researcher’s presence and were free to alter any agreed-upon timeframes. Regarding the use of interview data, participants were asked for their consent for anonymised quotations to be used, otherwise their words were paraphrased. Before the collaborative workshops, in the case of participants who had not provided written informed consent as part of earlier study activities, they were asked to provide verbal consent for workshop data to be used for research purposes, which was recorded in an Excel spreadsheet.

## 3. Results

Multimorbidity was a new term for many of our respondents. A familiar scene during interviews, for instance, was a participant pausing mid-conversation to search for the term: “multi…what do you call it?…bidity”. Familiarity and prior engagement with the term was most pronounced in the international research community, HIV programme, and professional healthcare networks, and continued to grow during the lifecycle of the study (not least because of our study’s influence). Those who were already acquainted with the concept generally understood it as the cooccurrence of multiple long-term conditions in one person, with HIV-NCD a commonly cited example. But what challenges and tensions the concept brought to the fore, its implications, and any reservations about prioritising multimorbidity were closely tied to participants’ particular positioning within the health system. In the following sections, we trace a narrative through the key themes that emerged through different ‘windows’ into multimorbidity including health seeking and delivery; policy and planning; medical training and professionalisation, and health data and research.

### 3.1. Whose Multimorbidity is “Better”?

> “Interviewee: Yes, my [younger] sister is asthmatic and diabetic as well, the one who comes after me is also asthmatic and diabetes, I am asthmatic too…We joke about our illnesses at times comparing who is better than the other, the one with HIV or diabetes plus asthma.

> Interviewer: Do you fear that you might have diabetes too?

> Interviewee: (laughs loudly) Yes, I am already asthmatic and HIV positive, I can just imagine being told I am now diabetic. Once you have diabetes the next thing is high blood pressure these two go together. Can you imagine all this burden on me? For now, I am okay without knowing, I will ignore the symptoms till I seriously get ill then I know it’s time.”

> (Patient_5, Mashonaland East, living with HIV, asthma, hypertension)

> “When I had HIV alone, I was confident that chances of living longer are high because I had known some people who were HIV positive for years. However, when I had diabetes, I felt robbed of life.”

> (Patient_14, Bulawayo, living with diabetes & HIV)

None of the PLWMM in our study had encountered the term ‘multimorbidity’ before our study. However, all talked comfortably about the ‘multiple’ nature of their conditions. As illustrated by the first respondent, they spoke about how different conditions interacted and were associated, the burden they exerted, and, indeed, which condition or combination of conditions were ‘better’ to have. Such appraisals, as the second respondent suggests, often foregrounded the differential experience of HIV and NCDs, reflecting the stark difference in the resourcing and organisation of services for these conditions. HIV+ PLWMM were, we found, comparatively satisfied with the quality of care for this particular condition. HIV prevention and treatment was decentralised and delivered by nurses, assisted by community health workers, near their homes, with only complicated cases needing referral/doctor support. Services were free, with consistently available medicine refills (which could also be collected at a community health post), integrated care for opportunistic infections related to HIV (e.g. TB), and often additional resources and support. In the higher-volume primary clinics especially in Harare, Bulawayo, and provincial centres, services were usually offered in a dedicated HIV clinic, referred to less explicitly as the ‘opportunistic infection’ or ‘OI’ clinic with their own separate staff, often supported by NGO-paid nurses.

The addition of NCDs or mental health conditions, however, dramatically shifted the experience of multimorbidity. Private clinics and certain “bougie” (i.e. high-resource) NGO-run HIV clinics were able to provide integrated, person-centred care for most NCDs, increasingly with an explicit multimorbidity focus. These one-stop-shop models were a common frame of reference among both PLWMM and health workers for what ‘good’ multimorbidity care for looks like:

> “What I learnt at [NGO-run HIV clinic] is that…The patient gets attended to as a whole under one roof, they don’t need to go elsewhere for other conditions, tests and collection of results like we do here. We need to upgrade and provide all services under one roof” (HCW_20, sister-in-charge, Bulawayo)

The nurse here was comparing partner-funded HIV-NCD care with the ‘standard’ of care for most living with NCD-related multimorbidity, which was provided through general clinic outpatient departments (OPD). OPDs were generally busy, overcrowded environments often staffed by only one extremely stressed, overworked and underpaid nurse. Typically this nurse was working with very little in the way of equipment, diagnostics, and medicines needed to perform the basic consultations per the EDLIZ guideline, with particular shortages noted with second-line NCD medications (<u>S2 Appendix</u>). Unlike HIV, user fees were required to visit urban clinics (with the exception of those <5 and >65 years old; rural clinics remain free), and all patients also had to pay for their medicines (either at the clinic or, if out of stock, at a private pharmacy). A further major bottleneck is that unlike HIV, for which most tasks have been shifted to nurses, initiation of treatment for most NCDs requires seeing a doctor – which, given the severe shortage of doctors, usually meant referral to a hospital clinic. Often, indeed, patients went straight to one of the central hospitals, either because they knew that they would not be helped at the primary level or because they were so ill that they required immediate hospital attention. While stable patients were referred to their local clinic OPD for ongoing (self-) management, HIV and NCDs would be managed separately. NCDs also normally required ongoing lifelong hospital visits – for quarterly reviews, any changes to medications, for any complications or further morbidities. All of this exerted a huge burden on PLWMM. While some felt that they were able to cope with cumulative costs, few were satisfied with the quality of care, and others felt unable to afford to ‘keep up’. They frequently had to make impossible choices about which condition to prioritize, which appointments to attend, or which medicines to buy. Ultimately the cumulative toll of health, social, and economic problems often inevitably led to secondary complications requiring more expensive inpatient care, as well as further conditions, frailty, and diminished coping capacity. Engaging with PLWMM and providers at different levels of care enabled us to sketch a care pathway for long-term conditions (Figure 2) that we used to engage further stakeholder groups.

**Figure 2.**
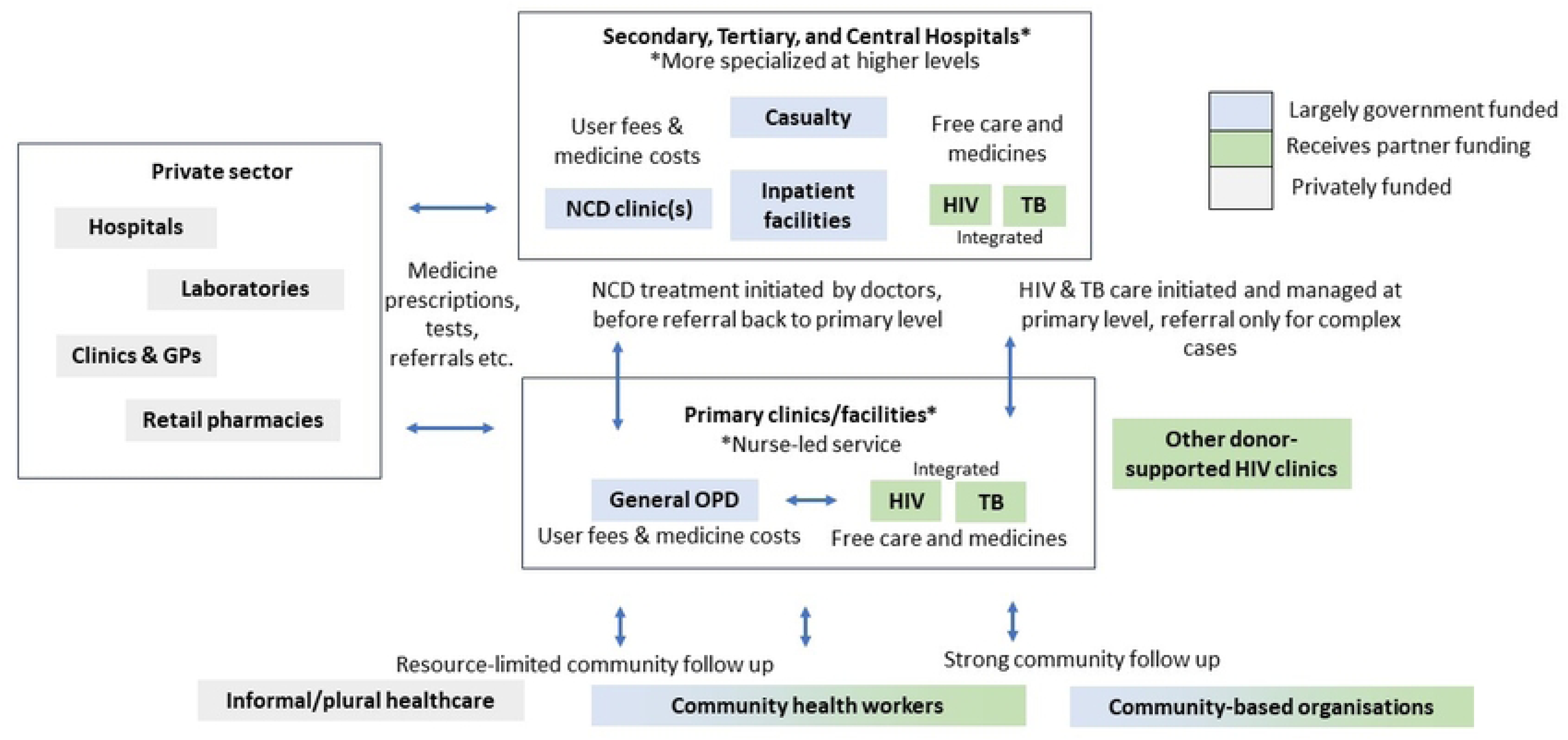
Illustrative care pathway for long-term conditions^#^. ^#^Considerable heterogeneity exists across provinces and districts, thus this pathway may not be representative of all referral possibilities

### 3.1 Siloes from “the Top” and the Erosion of Comprehensive Primary Care

When talking to policymakers and public health practitioners especially in Harare and Bulawayo about multimorbidity, one of the recurring narratives was that people are now living longer in part because of the successes of HIV care, resulting in a greater burden of NCDs and mental health conditions. Yet, Northern partners preferred to work through ringfenced funding for HIV, TB, and malaria which created ‘siloes’ within the health system. As one policymaker argued:

> “We have had HIV for quite some time, and the survivors for those living with HIV also have other conditions, in that same person you can find malaria, TB and HIV or other non-communicable diseases then it becomes imperative to probably shift. The initial design of these grants was to look at TB, HIV and malaria which created siloes within the health system and the move away from there is taking time”. (Policymaker_1, MoHCC, Harare)

Through interviews and participant-observation, we found that these fragmented funding streams have very real consequences in the day-to-day management of the health system that prevented working across different disease areas. Certain directorates, for instance, notably those relating to HIV programming, were large, well-funded and supported by lower-level implementing personnel at provincial, district, and facility levels. By contrast, departments without much in the way of donor backing, for instance those for NCDs and mental health directorates, were made up of a handful of individuals, with limited funds to strategize and plan, and far fewer focal persons at lower levels to support implementation. Against this backdrop, an air of competition and protectionism characterized inter-programme relations, likened by one policymaker to fighting over a “carcass” (Policymaker_2, Harare). This worked against collaboration between disease programmes and could result in separate, sometimes contradictory policies and interventions. This was even within areas that had historically been the focus of integration, for instance TB and HIV. Walking with another policymaker through one of the long corridors housing different disease-specific departments, they observed that:

> “The departments of NCDs, HIV, TB and malaria hardly meet to discuss patient issues, but they are all under the same roof which is a challenge already. At the higher level the focus is on the condition and as we go to the lower levels, they see a person and not a condition” (fieldnote excerpt, Policymaker_3, Harare)

Those at “lower levels” certainly had much to say on the matter. Health workers, facility managers and district/provincial decision-makers, particularly those who had experienced and contributed to the prior strength of the system in the early postcolonial era, were fierce critics of the current scenario. It was indeed, a provocation offered during a collaborative workshop that was thought by the assembled to have hit the nail on the head. Perplexed by why multimorbidity was being discussed like it was something new that had arrived once the HIV epidemic had stabilised, one provincial policymaker pointed out that the country’s primary clinics were already designed to be multimorbidity clinics. It was siloes from “the top” that were inhibiting the “already-integrated” nurse from delivering integrated care:

> “Already at the primary healthcare level you get [Bulawayo] clinic, it’s a multimorbidity clinic. You go to [Bulawayo] clinic, and it’s a multimorbidity clinic…Our priorities should be integration from the top. These siloes come to the already integrated primary care nurse who deals with all the conditions under one roof. Fund those clinics at the right places and let the proper pyramid in primary healthcare work as it used to be” (Policymaker_8, Bulawayo)

Responding to multimorbidity, for her, was a matter of allowing the lower levels of care to do what they were already designed and previously able to do in the 1980s and 1990s before the HIV epidemic.

Integrating primary care services and combatting entrenched ‘siloes’ was, on the one hand, explicitly written into National Health Strategy 2021-25.(30) This was being enacted through a number of promising initiatives including an early-stage pilot of the WHO Package of Essential Non-Communicable Disease (PEN) Programme (designed to decentralise the management of common NCDs), new NCD guidance within the latest HIV Operational and Service Delivery Manual (OSDM) on the integration of HIV-NCD services, and a recent application to the Global Fund to fund medicines for common NCDs among people living with HIV which was hoped to “unlock” previously inaccessible resources for NCDs: “I believe with integration there is a lot of resources that can be unlocked which can then spill over” (Policymaker_1, Harare). Many felt, however, that while these were positive steps, they would be insufficient to respond to multimorbidity and will likely only benefit HIV+ patients and buttress the HIV programme. The underlying issue, many felt, was that the international funders neither had the resources nor the inclination to take the step beyond vertical funding that would be needed to respond to multimorbidity. Nor, despite the emphasis in the National Health Strategy on combatting siloes, was reinvigorating comprehensive primary healthcare felt to be especially high on the government’s agenda either. For as several suggested, head office was preoccupied with developing specialist capacity at the quaternary level, which would predominantly benefit the country’s political and economic elite.

### 3.3. Clinical (Sub-)Specialism over Generalism and Public Health

This perceived drive towards higher-level, specialist medicine, relative to lower-level ‘generalist’ care was a key theme brought to the fore when discussing multimorbidity with clinicians and educators. Specialist trajectories were inscribed at multiple levels: from individual-level pressures to enhance earning potentials and opportunities; to institutional aspirations towards becoming recognised centres of excellence in research, training, and care; to national level strategies (e.g. “Education 5.0”) to enhance specialist training, biotechnology, and novel drug development. Speaking to one specialist-in-training (Academic_2, Harare) about their hospital’s mission statement of one hospital to become a superspecialist centre of excellence by 2030, she related that these aspirations were still a way off. While specialist training (e.g. internal medicine) was offered at the central teaching hospitals, sub-specialising (e.g. in cardiology, neurology, etc.) required advanced training in South Africa or overseas, a route very few were able to afford, less motivated to return to practice in Zimbabwe – fewer still in the public sector. For those working within the public sector, the ability to practice one’s (sub-)specialty was severely constrained by available diagnostics, treatments and, indeed, what the patient was able to afford. Referrals to the private sector – often, indeed, to one’s own private clinic – was often the only available route, but this was not an option in many cases. And, indeed, such was the need and resource scarcity that it was impossible to exclusively practice one’s speciality. The hospital OPDs and general medical wards were overflowing, many with multimorbidity, at least in part because of the lack of capacity at lower levels of care to treat these conditions or prevent them from needing specialist hospital-level attention. As a result, as one sub-specialist put it, “we [sub-specialists] are all general with an interest in one area.” (Academic_3, sub-specialist physician and lecturer, Harare).

If generalism was the reality even for the country’s top sub-specialists, it was certainly the case for the majority of physicians working at lower levels of care. The problem was that the development of generalist skillsets was, as several observed, not adequately valued or supported. Both classroom learning and rotations were taught around diseases and organ-systems – “in siloes”, as another lecturer described (Academic_1, sub-specialist physician and lecturer, Harare), which ran contrary to a multimorbidity approach. On the one hand, a common line of thinking was that trainees were given ample opportunity to consolidate specialist knowledge including in the management of multimorbidity during their internships, which often took place in district hospitals. At the same time, it was argued by others that this ‘on the job’ approach to ‘putting it all together’ was not felt to be sufficient, and reflected a lack of value placed on generalism compared to the advanced training expected of specialists. As one Bulawayo-based general practitioner argued:

> “A good generalist is someone who should be able to take care of the most common and prevalent conditions that burdens the local community that he practices in and should be in a position to be a team leader in the preventive aspect of medicine… Let’s try to prevent the conditions from getting worse to require specialist care… But we don’t do that, why, because people don’t know how important a generalist is, a family physician is, a primary care physician is.” (HCW_43, general practitioner, Bulawayo)

That new physicians were ‘default GPs’, as a number of physicians put it, was contrasted with high-income settings, such as the UK, where post-basic certification was required. While in the 1980s, there was a well-subscribed MMED in General Medicine similar to the UK model, this was discontinued in the 1990s. A more recently introduced MMED in Family Medicine remains undersubscribed, especially compared to the MMed Internal Medicine (for those looking to (sub-)specialise) and the MSc Public Health. Training in family medicine, we found, was disincentivised by the fact that graduates were, as ‘generalists’, still not regarded as (or able to charge as) ‘specialists’: “I think there is then need for a move to formalise some of these professions so that even if they do these kind of training they actually, well I will call them specialized in a way” (Policymaker_4, Matabeleland South). Those training in public health, meanwhile, often ended up transitioning towards either MoHCC head office, NGOs, or research institutions. This not only removed these skillsets from the clinical level but meant that they were unevenly absorbed into the better-funded programme areas.

The training of nurses and other cadres was, on the whole, perceived to be more ‘generalist’ in orientation. Indeed, the training of general nurses and village health workers has historically been one of the great strengths of Zimbabwe’s primary healthcare system, with the ‘old style nurse’ being adept not only in managing all common conditions found in the EDLIZ guideline but also really ‘thinking through’ your problems. Standards were felt to have dropped in recent years due to resource scarcity, staff attrition, and resulting a lack of trainers and mentors, often forcing some older nurses out of retirement. This is at precisely the time when, as the senior nurse above put it, the “already integrated primary care nurse” (Policymaker_6, Bulawayo) was being pulled in different directions by the increasingly fragmented needs of disease-specific programmes, disproportionately for HIV, TB, and malaria, each coming with additional evidence-based clinical practice guidelines, training packages and monitoring and evaluation (M&E) systems. In these there was, as one policymaker specialising in service integration put it, “no room for other diseases, or usually just a small component” (Policymaker_3, Harare). Such a ‘small component’ included the recent inclusion of training in carrying out the aforementioned expansion of OSDM guidance to include hypertension, diabetes, and depression. Meanwhile, there were no in-service training for NCD management beyond HIV, with most nurses relying on what they learned in basic training, evidenced in our facility survey by few nurses trained in the last 2 years in diabetes, hypertension, asthma, or depression (<u>S2 Appendix</u>). Together, falling standards and the fragmentation of training around disease-specific programmes was seen to work against the previously strong generalist capacities underpinning Zimbabwe’s nurse-led primary care, so important for managing complex health needs like multimorbidity at lower levels.

### 3.4. Fragmentation and Demands of Health Data

If tensions between specialist and generalist training came to the fore when considering multimorbidity from human resources perspective, the fragmentation and demands of health data presented another layer of complexity to the challenge. Health informaticians and data experts described for us a complex data ecosystem that made up the ‘back end’ of Zimbabwe’s health system, one that had become progressively more expansive and fragmented in recent years. It comprised patient books/cards and records; disease tally sheets (referred to as the “T-series”); a plethora of single-disease programme-specific registers; and the recent introduction of Electronic Health Records (EHR), intended to phase much of this data into a more integrated electronic format. It also includes the MoHCC central health information system, DHIS2 (a widely-used system across Africa), into which routine data sources are fed through structures of upward reporting from facility to national level.

In terms of clinical management and follow-up, tracing patients with multimorbidity was challenging because, while in theory patient-level records should include all known conditions about a patient, this was not always the case. Even at primary clinic level, there were separate books for the OI department (the so-called “green book”) and general OPD, often making it hard to keep track of HIV+ patients with NCDs and vice versa, particularly if the patient forgot their book(s). Facility-held records for long-term conditions included HIV-related and TB registers and the ‘chronic’ register for common NCDs (hypertension, diabetes, asthma, epilepsy, and depression). While facility-held, registers unlike the patient booklets were more condition-oriented than the latter, with the same patient often appearing across registers with no cross-reference, making it challenging to identify patients with multiple conditions. In some facilities, we found that nurses would informally record a patient’s other conditions against their name wherever it appeared, so that whoever attends to the patient knows they need to update for all other conditions:

> “If a patient’s clinic number is one on the HPT register and number 3 on DM register then on their book it will be recorded for example 1HPT and 3DM. It will show their clinic number in each condition they have. The clinic chronic register is all we have. If the patient is on ART his or her O.I number will be written on their booklet too”. (HCW_9, Acting sister-in-charge, Mashonaland East)

The challenge, however, once patients were sorted into a disease specific register or programme for ongoing chronic management of a particular disease, it was likely that a patient would be fast-tracked to registers for this particular disease in future consultations, and less likely, especially in a resource/time-constrained environment, that clinicians were optimally thinking about other conditions unless the patient explicitly mentioned them. On the one hand, EHR was felt by many to hold considerable promise for ameliorating fragmentation and improving the tracking of patients through a unique patient identifier, in theory across facilities and levels of care. Currently, however, EHR is still inconsistently available within and across provinces, often down due to connectivity or electricity issues, and remains currently only operable within the public sector. Perhaps more fundamentally, EHR is still ultimately an e-copy of the paper-based system, with disease ‘modules’ taking the place of physical registers. This does not actually depart from disease-specific health information and, in fact, may reinforce it. EHR, for instance, was initially funded by the HIV programme, whose module is thus more advanced than the ‘chronic’ module. There has also been funding provided from the other partner-backed programmes to have their modules developed and updated – much of which is occurring without necessarily considering of the overall integration of the system.

Data used beyond the immediate clinical encounter – for disease surveillance and control, annual health reporting, monitoring and evaluation (M&E) of specific programme targets, and policy and resource allocation – was, if anything, more single-disease focused. Abstracted from the tally sheets (T-series), programme registers, and other forms, data reported upwards were largely cross-sectional – basically counts of disease conditions rather than counts of diseases in a single person: “The moment you run an aggregated system certainly there is no way you would expect figures that relate to a single person” (Health_Infor_3_Policymaker_5, Harare). The data were also heavily biased towards the programme-backed diseases (Figure 4). The better-funded programmes, for instance, have additional, high-resource M&E infrastructure that runs alongside MoHCC standard reporting systems. It includes standardised programme-specific registers (corresponding to their clinical guidelines); finely-differentiated indicators; regular visits and back-and-forths on data quality at the facility level; and M&E officers and analysts working at sub-national level all the way upwards. Certainly at the facility level, the heterogeneity of programme documentation meant a considerable paper burden on the nurses, experienced as representatives from one programme coming one day with demands, the next another programme doing the same, who were often felt to have a distinct lack of knowledge of the reality on the ground or the overall impact of the paper burden was having. “It demoralises us!,” as one nurse from Matabeleland South exclaimed. Further obscuring challenges on the ground was that, while the facilities were encouraged to examine data to identify challenges and solutions, it was felt that rarely were these efforts reciprocated, leading to despondency in reporting what could be vital sources of information:

> “We used to sit religiously interrogating the data then we would come up with a list of challenges and proposals. We ended up copying and pasting because nothing was changing. I think these are the things that should be informing policy so that policy changes.” (HCW_42_Policymaker_7, District Medical Officer, Matabeleland South)

A further knock-on effect of the demands of health data was that, while some data was of high quality non-programme backed diseases, which had neither the standardised registers, nor the human resources to chase data quality, nor the analytic capacity, were far more likely to be incomplete and left behind. NCD data, in particular, were felt to be under-reported and of poor quality, with whatever data making it up to DHIS2 going there only to “die”:

> “Nobody reports NCDs! We’ve lagged for so long we’ve let things go, even DHIS2 for schizophrenia – that data is there but not consumed, so it dies.” (NGO_6, NGO physician, Bulawayo)

On the one hand, while still early days, there is recognition of the need to improve data and information systems for NCDs but also, increasingly, multimorbidity. In terms of NCDs, this includes recent efforts to improve M&E infrastructure for NCDs as part of the WHO PEN programme, as well as plans to more regularly conduct the WHO STEPS survey which includes a component on NCD risk factors (the last of which was conducted in 2006 and is still being used in the National Health Profile[33] and Strategy[34]). There is also diagnostic yields from NCD screening initiatives conducted by HIV implementing partners; routine data available from HIV donor clinics able to support fully integrated NCD care; and the recent addition of new data points in DHIS2 on certain HIV-NCD data points, corresponding to the expanded HIV-NCD integration guidance in the HIV guidelines. Finally, there a growing body of clinical, epidemiological, and social research on multimorbidity in Zimbabwe (the current project among them), and this research is starting to push the horizons beyond what has been a fairly HIV-centric discourse until fairly recently. Of note, however, is that the knowledge architectures within which researchers and programmers are starting to understand and develop interventions for multimorbidity remain constrained to a large extent by the infrastructure already in place centred around HIV and TB. Thus in our experience, when it comes to questions around where to start – where to integrate from, on what to model our interventions – answers tend to gravitate towards the known quantities of HIV and TB.

**Figure 3.**
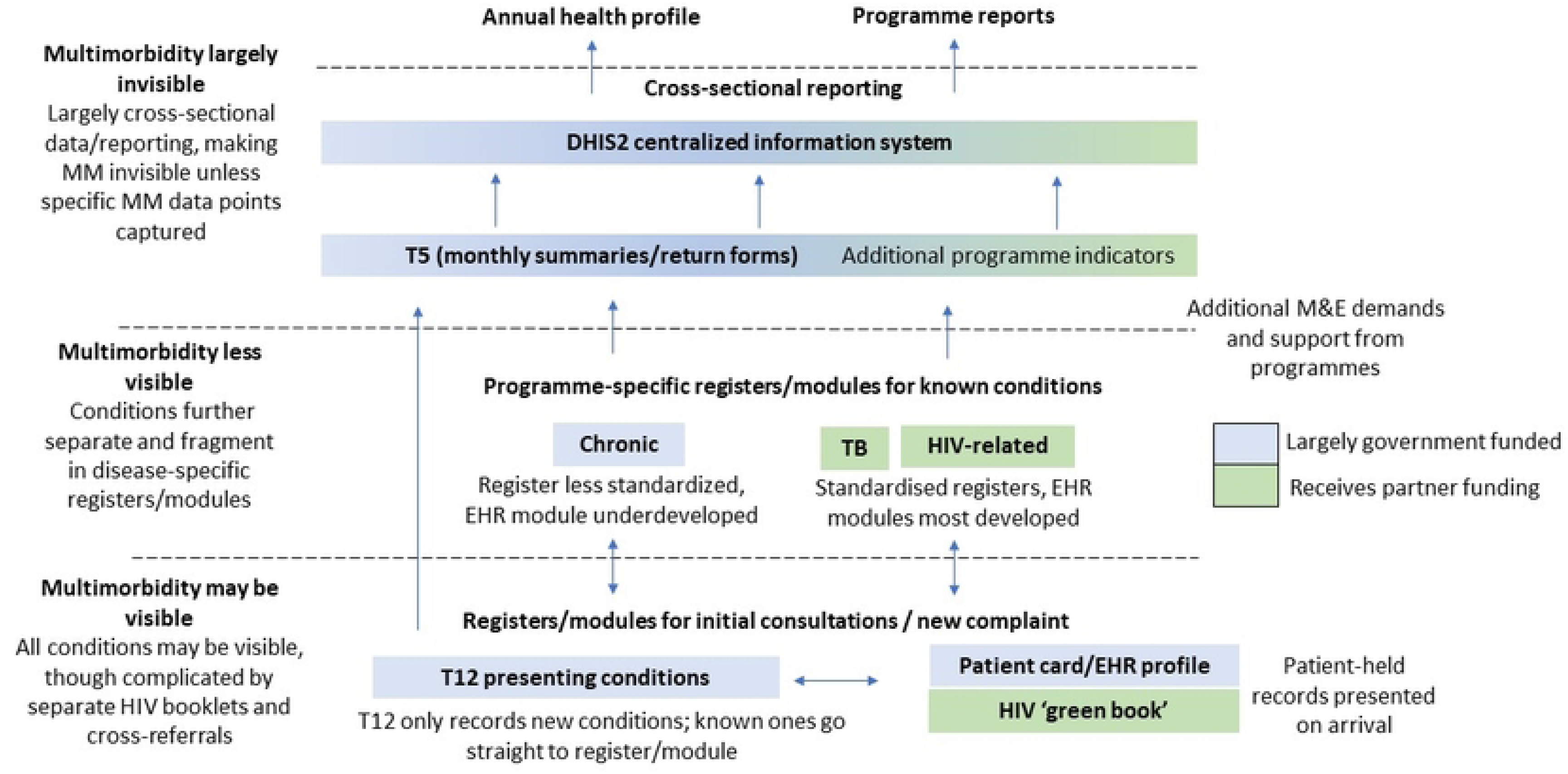
Data Flows for Long-Term Conditions and Multimorbidity.

## 4. Discussion

In this study, we used a participatory ethnographic research design to gauge and integrate different perspectives on the challenge multimorbidity in Zimbabwe. Bringing together different viewpoints at a time when multimorbidity was just emerging on the policy radar allowed us to produce a rich, holistic description to inform priority and agenda-setting while keeping sight of the possible limitations of elevating it as a specific priority. Overall, we found that multimorbidity almost unanimously resonated with respondents, who positioned multimorbidity as fundamental challenge to the current organisation of the health system. Multimorbidity was being most actively being engaged with from an HIV perspective and by the international research community, with conversations around multimorbidity beyond these groups somewhat less explicit or coordinated.

Our findings building upon a growing body of social research on multimorbidity in Africa. Consistent with findings in Malawi,[14] Ghana[35], Ethiopia,[15] and South Africa,[12,13] we found that, for PLWMM and their families, the concept of multimorbidity captured the tremendous health, economic and social burden imposed by the competing demands of different long-term conditions that were, in anything, exacerbated by the ‘vertical’ organisation and uneven resourcing of care around single diseases. The challenges faced by PLWMM, we went onto show, were a downstream reflection of wider tensions facing Zimbabwe’s policymakers and planners, for whom multimorbidity presented as profound managerial disenfranchisement characterised by contestation over the “carcasses” of disease-specific funding – a situation that rings all-too familiar with accounts of other African governments’ engagements with ‘global health’.[36–38] Such administrative challenges in turn speak a yet wider, well-described rupture in the biopolitical order from the promises of nation-state-led public healthcare systems following Alma Ata to the bankrupting of such systems and incursion of fragmented donor funding following HIV.[39] With Zimbabwe having made among the greatest strides towards comprehensive primary care grounded in its essential medicines programme,[25] the system’s downfall and current inability to even provide basic NCD medicines was experienced by many as a profound sense of loss of grassroots-level primary care. Further gutting the system is the strain placed on clinical and public health professionals being trained and socialised within the system, for whom multimorbidity presents a series of interlocking pressures towards specialised hospital medicine, the private sector, NGOs and research institutes. This corresponds with a weakening and devaluation of the strong generalist positions and skillsets that previously made comprehensive care possible at the lower levels.[17,40] Finally, health information and data, rather than revealing a problem with the status quo, functioned to legitimise it. From a health data perspective, multimorbidity manifested as the systematic ‘papering over’ of patient complexity through echo-chambers of disease-specific data capture, upward reporting, and feedback, with the ‘best’ data mediated by institutes and analysts in the global North.[41] This made it difficult to track multimorbidity patients over time and space; imposed a paper burden on nurses that detracted from time with patients,[17,42] while concurrently making the challenges faced at the care delivery level almost impossible to see, less listened and responded to, higher up. This demoralising situation functioned to disempower nurses, exacerbate the burden on patients, while reproducing the hypervisibility and prioritisation of already-funded diseases.

Multimorbidity may have been a meaningful and useful concept for most. However, the current direction from which most explicit multimorbidity discourse is coming, coupled with the perspectives of our respondents, gives reason to pause. Multimorbidity has been promised to lead us towards integrated person-centred care,[18] however as critics have pointed out, it has followed a familiar pattern within global health, focused on quantifying and objectifying it such that it can form the basis of new evidence-based interventions..[16] As this has been transposed into a global health problem, multimorbidity has found fertile ground in a ‘clashing epidemics’ narrative[17,43] that, in Zimbabwe as elsewhere,[44] has been readily absorbed within the HIV programme’s remit, reflected in growing knowledge of HIV-NCD comorbidity, progress with integrating certain NCDs into the existing HIV structures, and plans to use HIV as a model for NCDs and multimorbidity beyond the HIV+ population.[45] But ethnographic evidence, both in high-income[18] and lower-income settings,[12] suggests that expanding or merging vertical programmes will not be sufficient to respond to the challenge of multimorbidity.[16,17] In the UK, Lynch et al. found that multimorbidity has failed to live up to its promise to deliver on person-centred care, partly because the emphasis has been on building bridges between an ever-growing number of (sub-)speciality areas, with very few talking about the *repair* or *restoration* of the more comprehensive primary care services that were in place prior to greater specialism.[18] In Zimbabwe, the picture was somewhat different. For many participants, Zimbabwe was already prepared for multimorbidity before HIV was extracted, given its own clinic, and made into something special. As we have also shown in prior ethnographic research, Zimbabwe’s historically strong, comprehensive primary care system remained a powerful memory and guiding frame,[25] and thus what it meant to respond to multimorbidity, for many, was at least as much about restoring or repairing what was within the system, as much as preparing ourselves for a ‘new’ emerging pandemic.[1]

Accordingly, the formative agenda that emerged from the collaborative interpretations of the study’s findings during workshops was oriented towards what it would take to restore the centrality of comprehensive primary healthcare and support the nurse-patient encounter (<u>S4 Appendix</u>). While the priority areas within the agenda are expansive, two topics occupied a great deal of the conversations and cut across several of the priority areas. One was the question of what integrated care meant for existing HIV and ‘OI’ infrastructure. Such conversations drew from other countries’ experiences with addressing this situation, including South Africa’s ICDM model,[46–49] INTE-AFRICA study in Tanzania and Uganda,[50] and Malawi’s Integrated Chronic Care Clinics,[51,52] the latter of which MoHCC representatives had visited recently. A common feature of such approaches is the reorganisation of care from an ‘HIV vs. all other conditions’ model to an ‘acute vs. (stable) chronic conditions’ model in which existing HIV clinic/spaces are repurposed such that all chronic patients, regardless of HIV status, are seen within the same clinic/space. Proposed benefits include: creating a platform for the decentralisation of chronic conditions previously treated at higher levels; more efficient use of clinic space and resources; more patient-centred management plans, guidelines, and training; a harmonised and less burdensome M&E infrastructure; and the destigmatisation of HIV;[48,51,53] – though to date, such efforts remain few in example, early stage, and with challenges of uptake, sustainability, and scale-up.[44,54] The Zimbabwe MoHCC and technical partners have recently participated in cross-country discussions exploring possibilities for such a model in this setting,[53] and, for the most part, the collaborator group in this study (some of whom took part in that initiative) welcomed such a model. At the same time, consistent with our primary data, a particular proviso was that another parallel silo is not inadvertently created, which may come from the reification the chronic clinic concept without consideration of what was actually needed in different settings and levels of care (e.g. in a rural clinic set-up where there was no ‘OI’ setup to begin with). Insofar as such model could be useful, it would be one focused not on taking chronic disease or multimorbidity *out* but putting HIV back *in*, leveraging resources and infrastructure where needed but only insofar as it helped restore and strengthen the generalist orientation and referral pyramid.

If chronic clinics occupied conversations at the service delivery level, a second cross-cutting conversation was the more upstream enabling environment needed to facilitate meaningful and sustainable change at the service delivery level. Enacting a multimorbidity-responsive health system, it was felt, required being able to draw together knowledge and experience across different academic and applied spheres and to really *listen* to experience on the ground – currently so difficult within a siloed, top-down environment in which priorities and pathways for action are determined by external funding, agendas, and analysts. Certainly a lot of this boiled down to the structure of health financing and resulting fragmented policy structures, and the first step was recognised as being stronger with external partners on the pooled financing mechanisms that do exist but are consistently been undercut by earmarking of donor funds to particular conditions.[34] At the same time, it was also recognised that decades of under-resourcing, disenfranchisement, decapacitation, and attrition have meant a ‘culture’ that is simply not conducive to knowledge co-production, collective problem-solving, or innovation. Recent work within health policy and systems research (HPSR) has stressed the importance of active investment in learning as a core pillar of health system strengthening, over more short-term wins that are often prioritised.[55,56] Part of this means enhancing domestic information systems, research, and analytic capacity, particularly to lessen the dependence on parallel extractive data systems. However, as our respondents made clear, data can only take us so far and easily become overemphasised. Just as important within a learning framework is elevating other modes of learning that transgress traditional knowledge-practice binaries: inclusive deliberative platforms, practical or ‘experiential’ learning, and ‘embedded’ research, oriented towards what Gilson et al. have referred to as “collective sensemaking”.[57,58] A learning health system approach, we suggest, may bring together of these more upstream imperatives expressed by our respondents, providing a quite radical but tangible framework for how knowledge, policy, and practice might synergise within a multimorbidity-responsive health system. Unless we explicitly invest in strengthening these more ‘upstream’ capacities,[16] then integrated or person-centred care models at the service delivery level – the dominant focus of evidence-based interventions – may be undercut by familiar systemic challenges.

Striving towards a ‘multimorbidity-learning’ health system may seem aspirational, particularly in the context of Zimbabwe’s challenging social, political and economic environment. However, the need for bold movements to challenge the status quo is urgent. COVID-19, while underscoring the need to take multimorbidity seriously, also made evident the waning international capacity for supporting lower-income countries beyond securitised public health measures.[59] Donor funding for HIV is expected to reduce drastically by 2030 as part of the transition towards a ‘maintenance’ model. Against this backdrop, health systems across Africa may soon need to have developed domestic capacity to respond to increasingly complex needs of both those living with and without HIV, while navigating an increasingly securitised global health agenda that may not be responsive to such needs.[59] At the same time, these developments and the potential loosening of ‘vertical’ programming may yet offer an opportunity to further the decolonising health agenda.[60,61] At this moment when multimorbidity seems to be reigniting calls for more holistic, person-centred approaches to medicine and global health in a way the primary care and NCD agendas have largely failed to do,[62] we suggest that this could yet be a fruitful construct (if framed carefully) for building and pooling resources toward a shared vision for the future of African health systems. This could be grounded in recognition of not only shared challenges in the current moment but also aspirations towards – or in Zimbabwe’s case, back towards – more integrated, adaptive, and ultimately more person-centred healthcare systems.

## Conclusions

In this article, we have sought to piece together the challenge of multimorbidity from the perspective of multiple actors across Zimbabwe’s health system. Our particular message has been that how multimorbidity is presented as a challenge, and by whom, matters. Multimorbidity, it is hardly new to say, is hoped to bring about a return to more holistic, upstream, person-centred approaches. But while it may be tempting to frame it as new and pressing, worthy of funding and attention amidst competing priorities, doing so may perpetuate the same challenges it is hoped to overcome.[16,17] Following other ethnographic approaches, we have proposed multimorbidity is pointed in a somewhat different direction, focusing on repair and restoration of older systems as an integral part of responding to what is (apparently) new.[18] Of course, it is not simple to turn back the tide, and there is need to consider other countries’ experiences with integrating care or otherwise putting systems in place in which HIV is not the centre. In doing so, there is need to expand beyond the care delivery, as the violence of vertical programming has impacted all aspects of health system functioning.[17] The learning health system, unsettling many of the knowledge siloes and hierarchies that continue to perpetuate single-disease thinking, may offer a framework for intervening at this level and thus towards more sustainability person-centred health systems.

## Data Availability

Data from the study has been deposited with the United Kingdom Data Service (UKDS). We have included all transcripts (in-depth interviews), health facility survey data, and second-order summaries of fieldnotes from observational work. In line with our ethical approval, we have excluded raw fieldnotes, workshop discussions, and audio-visual diaries due to the challenges in maintaining confidentiality. The DOI for the data in UKDS is: https://doi.org/10.5255/UKDA-SN-857310.

## Acknowledgements

We thank the Zimbabwe Ministry of Health and Child Care (MoHCC), Provincial and District authorities in Harare, Bulawayo, Mashonaland East, and Matabeleland South, the University of Zimbabwe and the National University of Science and Technology for their invaluable guidance, support and collaboration. We express our gratitude to all the participants who took part in the study, whose experiences, insights, expertise and often collegiality were invaluable to the study’s findings. We also thank all administrative and technical staff who helped to ensure the operational success of the study, including the considerable work that went into organising and implementing the collaborative workshops.

## Supporting Information

**S1 Table. Breakdown of participant demographics of people living with multimorbidity (PLWMM).** Provides information about the demographics of PLWMM included in the study, including age, sex, province, and conditions.

**S2 Table. Availability of medicines, equipment, diagnostics, and training for common NCDs in 30 health facilities.** Provides select quantitative findings from the health facility survey regarding medicines, equipment, and training for NCDs.

**S3. Table. Priorities and key institutions for responding to multimorbidity in Zimbabwe.** Presents the main priorities and focal institutions for responding to multimorbidity identified during the final collaborative workshop held in Harare 1^st^ December 2023.

**S4 Checklist. COREQ Checklist:** Includes a completed COREQ checklist which is commonly used for reporting on qualitative studies. Domains include: research team and reflexivity; study design; and analysis and findings.

